# Sickle-trait hemoglobin does not influence *Anopheles* biting rates

**DOI:** 10.1101/2025.11.15.25340300

**Authors:** Christine F Markwalter, Emmah Kimachas, Erastus Kirwa, Joseph Kipkoech, Samuel Kahindi, Lucy Abel, Zay Yar Han, Judith N Mangeni, Andrew A Obala, Wendy Prudhomme O’Meara, Steve M Taylor

**Author notes:** These authors contributed equally to this work (Emmah Kimachas, Erastus Kirwa).

## Abstract

Children with sickle cell trait (HbAS) are protected against severe and symptomatic *Plasmodium falciparum* malaria. While several within-host resistance mechanisms have been investigated, it is unknown whether this protection may be attributable in part to reductions in exposure to *P. falciparum* parasites via mosquito bites. In a 15-month cohort in Western Kenya, we matched mosquito bloodmeals to human hosts based on short tandem repeat (STR) genotypes to determine individual mosquito biting rates. Using a multilevel multivariable model, we assessed mosquito biting behavior with respect to human β-globin genotypes and found no significant difference in the biting rates between individuals with HbAA and HbAS genotypes (biting rate ratio (BRR): 1.23, 95% CI: 0.86 - 1.77). These findings suggest that protection from malaria conferred by sickle trait is likely not attributable to reduced exposure to infectious mosquito bites.

**Author Summary:** Sickle cell trait (HbAS) is protective against severe and symptomatic malaria. Here, we investigate whether β-globin genotype (HbAS vs HbAA) is associated with differential mosquito biting rates. In a 15-month longitudinal cohort study in Western Kenya, we matched blood meals to community members based on short tandem repeat genotyping. We found no difference in biting rates across human β-globin types, suggesting that protection from severe and symptomatic malaria conferred by sickle trait is not attributable to reduced exposure to infectious bites.

## Introduction

Sickle cell trait (HbAS) protects individuals, especially children, from severe and symptomatic malaria (1). Several mechanisms for HbAS-mediated protection have been proposed, including differential acquired (2) and innate immunity (3), enhanced parasite clearance (4), and reduced cytoadhesion (5,6). However, it is unknown whether this protection may be attributable in part to reductions in exposure to *P. falciparum* parasites.

Exposure to *Plasmodium* parasites depends on the frequency of mosquito-human interactions, which are influenced by host factors including odor, body temperature, and carbon dioxide (7). Human blood characteristics may also influence attractiveness to mosquitoes. *Anopheles gambiae* fed more frequently on individuals with blood group O compared to other ABO types (8,9). On artificial feeding experiments, *An. stephensi* preferred blood group AB, followed by A, B, and O (10). The beta-globin variants hemoglobin S (HbS) and C (HbC) have been associated with increased transmission of *P. falciparum* from humans to Anopheline vectors (11), but it is not clear if this results in part from increased contact with vectors. To date, no studies have explored whether *Anopheles* mosquito biting preference is affected by the human host’s beta-globin type.

Here, we examine whether β-globin genotype (HbAS vs HbAA) influences mosquito biting preferences. In a 15-month longitudinal cohort study in Western Kenya, we matched 516 blood meals to community members based on short tandem repeat genotyping. We hypothesized that *Anopheles* biting rates may be associated with sickle cell trait.

## Results

The analytic dataset consisted of 52 households across 4 villages from July 2020 to September 2021 in which 313 individuals with HbAA or HbAS were at risk of being bitten. In this context, we collected 2841 female *Anopheles*, of which 1491 were freshly fed. Out of 890 bloodmeals that were STR typed, 621 returned human alleles and 516 matched to individuals in the cohort, comprising 545 biting events. We also collected 3677 DBS, of which 910 (25%) were positive for *P. falciparum*.

**Table 1.** Characteristics of individuals with HbAA and HbAS. The individual with HbSS has been omitted from this table.

| Characteristic | AA N = 244<sup>1</sup> | AS N = 69<sup>1</sup> | p-value<sup>2</sup> |
| --- | --- | --- | --- |
| Age |  |  | 0.5 |
| <5 | 32 (13%) | 6 (8.7%) |  |
| 5-15 | 86 (35%) | 28 (41%) |  |
| >15 | 126 (52%) | 35 (51%) |  |
| Gender |  |  | 0.8 |
| Female | 124 (51%) | 34 (49%) |  |
| Male | 120 (49%) | 35 (51%) |  |
| Proportion of time slept under net<sup>3</sup> | 0.75 (0.33, 1.00) | 0.60 (0.27, 1.00) | 0.12 |
| Proportion of time infected with *P. falciparum*<sup>3</sup> | 0.21 (0.14, 0.33) | 0.27 (0.14, 0.38) | 0.059 |

<sup>1</sup>n (%); Median (Q1, Q3) <sup>2</sup>Pearson's Chi-squared test; Wilcoxon rank sum test <sup>3</sup>Proportion of time based on monthly sampling

Among the 313 people in this study, 244 (78%) had HbAA, 63 HbAS (22%). There were no significant differences in gender or age distributions between β-globin genotypes. Similarly, there were no statistical differences in the proportion of time infected with *P. falciparum* or the proportion of time slept under a net across β-globin types.

We estimated the monthly biting rates for each individual based on the observed number of bites received and time at risk over the 15 month study period. In both groups, biting was highly overdispersed; the median estimated bites per month was 0 for HbAA and 1 for HbAS, and mean bites per month were 2.4 (HbAA; sd: 5.7) and 3.0 (HbAS; sd: 5.5) Among individuals who received at least one bite, the median number of bites per month was 2.3 for HbAA (IQR: 1.2 - 5.8) and 3.5 for HbAS (IQR: 1.4 - 7.3), and this difference was not statistically significant (Wilcoxon rank-sum, p = 0.094) (**Fig 1**).

**Fig 1.**
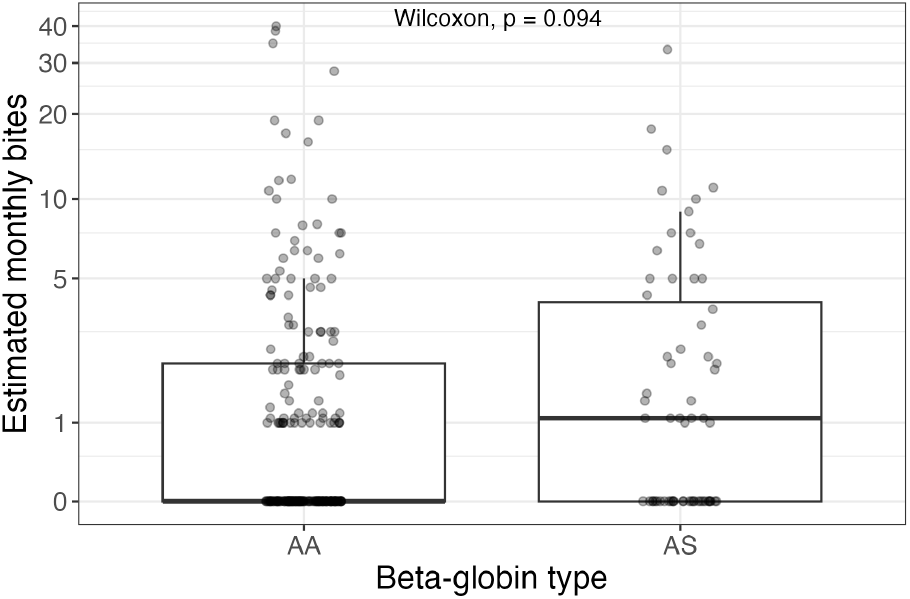
Estimated monthly bites per person by β-globin type. Each point represents one cohort member. Note the y-axis is log-transformed to visualize biting rates between 0 and 10 bites per month.

In a multilevel multivariable model including 1330 person-nights at risk, relative to people with HbAA, people with HbAS had similar mosquito biting rates (Biting rate ratio (BRR) 1.23, 95% CI: 0.86–1.77) (**Fig 2**). Consistent with previous observations, higher biting rates were observed among males (relative to females; BRR= 1.64, 95% CI: 1.19–2.26), children aged 5–15 years (relative to adults; BRR 1.43, 95% CI: 1.02—2), and participants with blood-stage infection (relative to uninfected; BRR 1.34, 95% CI:1.04–1.71); lower biting rates were observed for people who reported sleeping under a net (BRR 0.49, 95% CI: 0.37 - 0.65).

**Fig 2.**
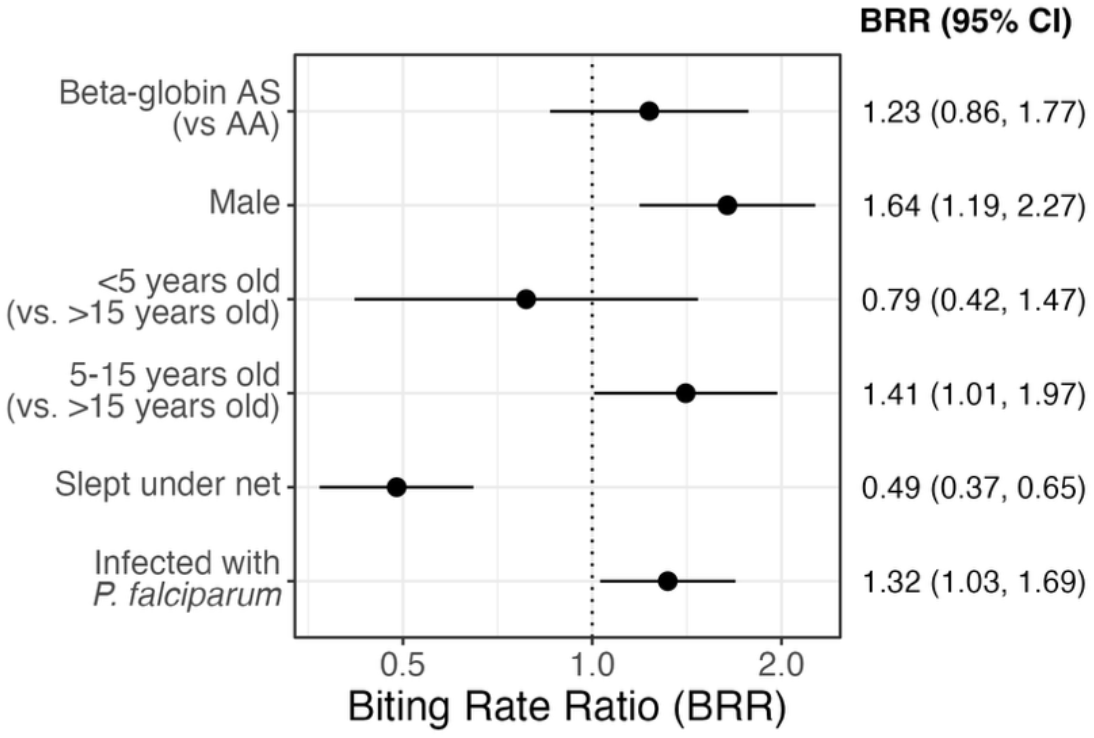
Risk factors for receiving female *Anopheles* bites. Biting rate ratios (points) and 95% confidence intervals for covariates in risk factor analysis based on 1325 person-nights.

Given that children under 5 suffer the highest incidence of malaria and benefit most from sickle-trait protection, we next explored whether age modified the effect of β-globin genotype on mosquito biting rates. The distribution of estimated mosquito bites per month stratified by age group (<5 years, 5–15 years, >15 years) and β-globin genotype (AA vs. AS) (**Fig 3A)** shows that the mean values (black bars) did not differ substantially between the two genotypes across any age category. Similarly, age-stratified models showed no differences in biting rate ratios for AS versus AA genotypes across age groups (**Fig 3B**); however small sample sizes, particularly in the youngest age group led to imprecise estimates.

**Fig 3.**
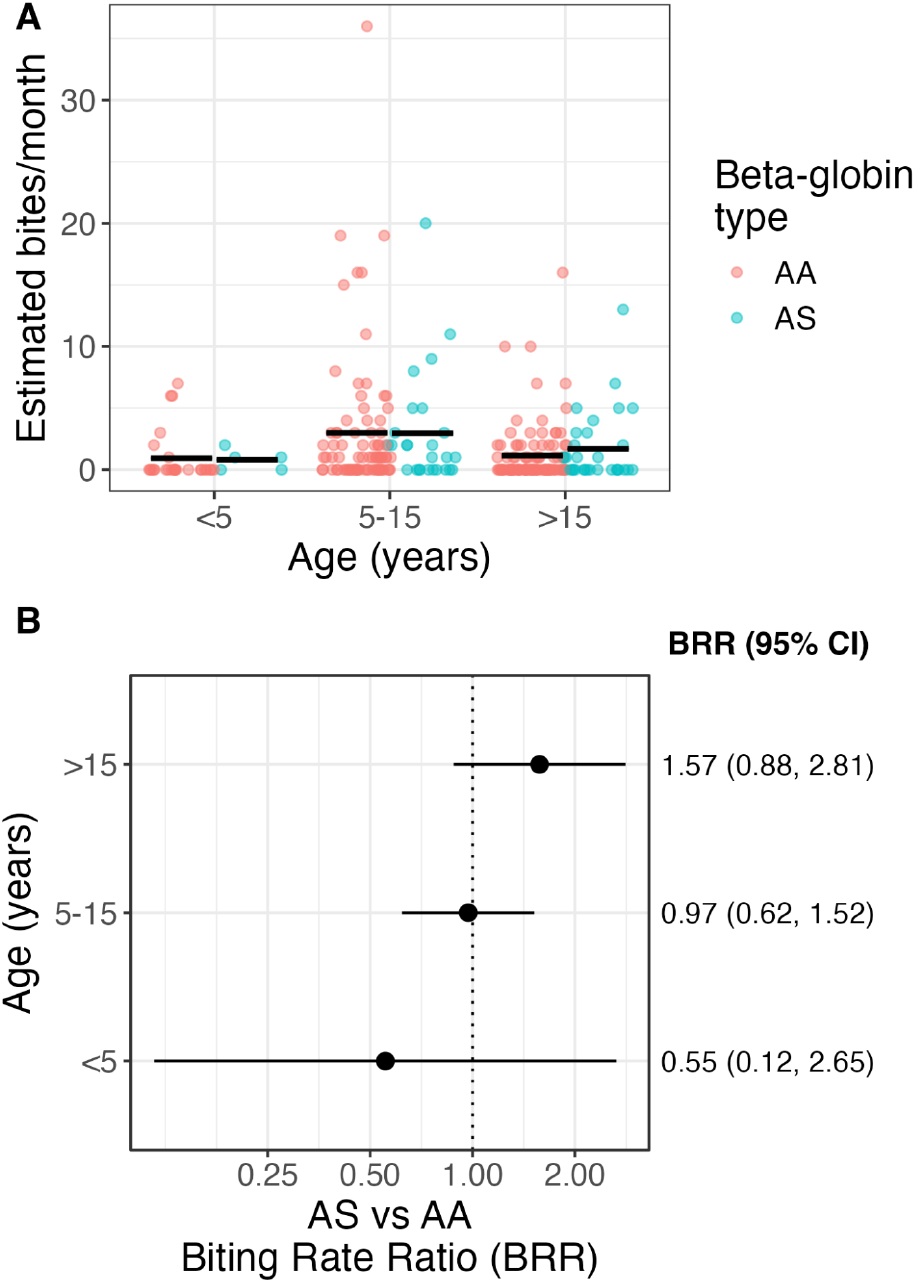
Female Anopheles biting rates by age group and β-globin type. A) Individual estimated monthly bites received, stratified by age group and β-globin type (color). Black bar represents mean monthly bites. B) Biting rate ratios (points) and 95% confidence intervals for HbAS vs HbAA, stratified by age category.

## Discussion

This analysis investigated whether sickle-trait hemoglobin influences mosquito biting preference in a malaria-endemic setting in western Kenya with a moderately high prevalence of HbAs of 22%. Over 15 months in 52 households, we detected human gDNA in 621 bloodfed *Anopheles* mosquitoes, 516 of which matched to cohort participants. We observed no significant difference in biting rates between people with HbAA and HbAS, indicating that sickle-trait hemoglobin does not modify the risk of being bitten by *Anopheline* mosquitos.

HbAS confers consistent protection against malaria by mechanisms that are incompletely understood, and therefore we explored the possibility that protection could be mediated, in part, simply by reduced biting by malaria vectors. The premise that biting rates may differ between HbAA and HbAS individuals is based on previous reports of differential biting across blood group antigens (8–10,12). The mechanism of this is obscure, but the expression of ABO antigens on a wide variety of tissues including endothelial and epithelial cells renders them legible to feeding mosquitos. In contrast, nonerythroid expression of beta-globin is rare (13), and therefore may be less detectable to Anophelines. Indeed, overall biting rates were not different between people with HbAA and HbAS, and nor did we observe reduced biting in age-specific groups, including in children who both suffer the highest incidence of malaria and benefit most from sickle-trait protection. Assuming similar overall biting rates also reflects similar biting rates by infected mosquitos, our results indicate that protection is not mediated by a reduced Anopheline biting of people with HbAS.

The observation that biting of people HbAS is not enhanced is relevant to the understanding of how people with sickle-trait participate in onward transmission. HbAS does not protect from blood-stage parasitization, and prior studies have demonstrated that individuals with HbAS who are infected with *P. falciparum* have enhanced transmissibility (11) compared to individuals with HbAA. Though this effect has been attributed primarily to elevated prevalences and densities of transmissible gametocyte forms (3,14) and the subsequent increased likelihood of transmission to mosquito during a feed (15,16), this increased transmission could also be enhanced if HbAS enhanced human-vector contacts. Though higher gametocyte prevalences in people with HbAS may drive *Anopheles* host preference and increased biting rates as reported (17), our results suggest that HbAS itself does not directly enhance vector biting.

This study has limitations. The small number of children under 5 years in our study (n = 38), which has limited our ability to precisely estimate biting rate ratios specific to this age group, which suffers both the highest burden malaria and the greatest protection from disease. Second, the captured mosquitoes represent only a small sample of all mosquitoes that feed within the community households; however, given the consistent collections, it is likely not biased toward biting a specific group of participants. Finally, the approach of matching bloodmeals measures successful mosquito feeds does not capture mosquito bites without successful feeding, which may contribute to disease risk.

In conclusion, we observed no difference in mosquito biting rates between people with HbAS and HbAA across all age groups. This suggests that protection from malaria conferred by sickle trait is likely not attributable to reduced exposure to infectious mosquito bites.

## Materials and Methods

### Ethical statement

We obtained written informed consent from all adult participants and from parents/guardians for participants <18 years. Verbal assent was obtained for children 8 - 17 years. Ethical approval was granted by the research ethics boards of Moi (572/2023) and Duke (Pro00113485) Universities.

### Study population and sample collection

This analysis is a subset of an ongoing longitudinal, community-based cohort study of up to 75 households across 5 villages in Western Kenya-Bungoma county (18). For the parent cohort study, *Anopheles* mosquitoes were collected twice per month within the cohort households by mechanical aspiration. Female *Anopheles* mosquitoes were dissected into head, wings and abdomen. Freshly bloodfed abdomens were pressed onto filter paper. Finger-prick dried blood spots (DBS) were collected monthly. We also recorded bed net use and symptomatic malaria episodes. From this cohort, this analysis included people in 4 villages who had consented to beta-globin typing and events observed during a 15-month period in 2020/2021 during which blood-fed mosquitos were matched to hosts, as described below.

### Molecular methods

#### Detection of *P. falciparum* by qPCR

This laboratory analysis has been published (18). Briefly, gDNA was extracted using the Chelex-100 from human DBS, head-thoraces, and abdomens of mosquitoes. gDNA from DBS and mosquitoes were acquired in duplicate using TaqMan real-time PCR assay targeting the *P. falciparum* pfr364 motif as previously described (19). Positivity was determined based on Ct values for the replicates, and parasite densities were estimated using the standard curves.

#### Human genotyping

Genotyping by short-tandem repeat analysis has been published (18). Each participant’s DBS and every mosquito abdomen identified as bloodfed and for which a blood spot was present were used for genotyping human blood using the Promega Geneprint 10 assay (20). We genotyped each sample type after extracting gDNA using Chelex-100. This assay did not amplify blood sources that are not human.

#### β-globin genotyping

gDNA from DBS from each participant was genotyped for HbS using a TaqMan SNP genotyping assay targeting the *rs334* variant that encodes HbS. We used TaqMan SNP genotyping assay C_1888768814_10 and amplified 1uL of gDNA template in a 5uL reaction using TaqPath ProAmp Master Mix, as directed. All reactions were run in duplicate on a Quantstudio5 machine on plates including as controls templates harboring known HbAA, HbAS, and HbSS.

### Bloodmeal matching

Bloodmeal matches from (18) on individuals who had consented to beta-globin typing were used in this analysis. Briefly, STR profiles in cohort members and mosquito blood meals were matched using the bistro R package (v 0.2.0) (20). The package enables matching for incomplete STR profiles and multisource bloodmeals; weight-of-evidence likelihood ratios (LRs) are calculated for every mosquito-human pair, and matches are determined based on thresholds for each individual mosquito. We then calculated the number of monthly bites on a participant: *Monthly bites* = *b*/(2*t*) × 30 *days*/*month* Where *b* is the total number of bites observed, and *t* is time at risk (in months) based on the number of monthly surveys. Thus, *b/2t* is the estimated nightly biting rate, given that mosquito collections were performed twice monthly.

### Risk factor analysis

We used a negative binomial regression to investigate the association between mosquito biting rates and β-globin genotypes (HbAA versus HbAS). The outcome variable was the number of mosquito bites matched to individual participants on a given night, and covariates included demographic factors (age, gender), use of bed nets (determined from the most recent monthly survey before the date of a matched bite; missing data was filled in using the monthly survey after the bite), and *P. falciparum* infection status, determined by qPCR at the nearest monthly sampling (within -30 to +7 days of the bite, except if RDT+ -14 to 0 days from bite then negative after receiving treatment). The model included a random intercept at the person-level and was adjusted for i) transmission season (high season March to August), ii) number of STR-typed mosquitoes in the household, iii) number of household members, and iv) number of people in the sleeping space.

### Data analysis and visualizations

All analyses and visualizations were performed in RStudio (v 2024.04.2+764) (21) with R v 4.3.1 (22) using the following packages: tidyverse (v 2.0.0) (23), ggpubr (v 0.6.0) (24), glmmTMB (v 1.1.8) (25), broom.mixed (v 0.2.9.4) (26), DescTools (v0.99.50) (27), and modelr (v 0.1.11) (28).

## Data Availability

Data from human participants in this study are not made available in an open repository due to privacy issues and conditions of IRB approval. Investigators interested in the dataset should submit a request to the Principal Investigators (W.P.O. and S.M.T.) and provide a brief study description/analysis plan. No identifying information will be shared, and data recipients will not be permitted to share data with other investigators.

https://github.com/malaria-house/trait_biting

## Data and code availability

Data from human participants in this study are not made available in an open repository due to privacy issues and conditions of IRB approval. Investigators interested in the dataset should submit a request to the Principal Investigators (O’Meara and Taylor) and provide a brief study description/analysis plan. No identifying information will be shared, and data recipients will not be permitted to share data with other investigators. All code that supports analyses and figures are available on GitHub: https://github.com/malaria-house/trait_biting

## Acknowledgements

We thank field technicians Ibrahim Khaoya, Lucas Marango, Ezna Mukeli, Eric Nalianya, Jane Nyongesa, Lilian Nukewa, Edith Wamalwa, and Aggrey Wekesa for their engagement with the study participants; Sarah Laing, Julius Maiyo and Emily Robie for operational assistance and coordination; Thynn Thane, Jillian Grassia, Jenna Decurzio, Daja Gatson, and Scott Langdon for sample processing; and Jamie Mills, Robert Rono, Francis Kithuku, Nikita Poujai and Heather Hille for administrative support. Ultimately, we are indebted to the study household members for their participation in this study. This work was supported by NIAID (R01AI146849 and R01AI179141 to W.P.O. and S.M.T. and K01AI175527 to C.F.M.).

## Notes

### Competing Interest Statement

The authors have declared no competing interest.

### Author Declarations

Ethical approval was granted by the research ethics boards of Moi University (572/2023) and Duke University (Pro00113485).

